# Synaptic oligomeric tau in Alzheimer’s disease – a potential culprit in the spread of tau pathology through the brain

**DOI:** 10.1101/2023.03.15.23287268

**Authors:** Martí Colom-Cadena, Caitlin Davies, Sònia Sirisi, Ji-Eun Lee, Elizabeth Simzer, Makis Tzioras, Marta Querol-Vilaseca, Érika Sánchez-Aced, Ya Yin Chang, Kris Holt, Robert McGeachan, Jamie Rose, Jane Tulloch, Lewis Wilkins, Colin Smith, Teodora Andrian, Olivia Belbin, Sílvia Pujals, Mathew H. Horrocks, Alberto Lleó, Tara Spires-Jones

## Abstract

In Alzheimer’s disease (AD), fibrillar tau pathology accumulates and spreads through the brain and synapses are lost. Evidence from mouse models indicates that tau spreads trans-synaptically from pre- to postsynapses and that oligomeric tau is synaptotoxic, but data on synaptic tau in human brain is scarce. Here we used sub-diffraction-limit microscopy to study synaptic tau accumulation in post-mortem temporal and occipital cortices of human AD and control donors. Oligomeric tau is present in both pre- and postsynaptic terminals even in areas without abundant fibrillar tau deposition. Further, there is a higher proportion of oligomeric tau compared to phosphorylated or misfolded tau found at synaptic terminals. These data suggest that accumulation of oligomeric tau in synapses is an early event in disease pathogenesis, and that tau pathology may progress through the brain via trans-synaptic spread in human disease. Thus, specifically reducing oligomeric tau at synapses may be a promising therapeutic strategy for AD.

## Introduction

Of the neuropathological hallmarks of Alzheimer’s Disease (AD), synapse loss is the strongest correlate of cognitive decline ^1–3^, and progression of tau pathology through the AD brain closely correlates with synaptic loss and cognitive symptoms ^4, 5^. Tau protein binds synaptic vesicles and is thought to have physiological roles in synapses ^6^. In animal and neuronal culture model systems, accumulation of hyperphosphorylated and misfolded tau within synapses causes synaptic dysfunction and synapse loss ^7–13^.

While neurofibrillary tangles (NFT) were historically considered neurotoxic lesions, data over the past decades indicate that the tau fibrils in tangles are relatively inert and that soluble oligomeric forms of tau are more toxic to synapses and neurons in mouse models of tauopathy ^14, 15^. Much less is known about synaptic tau and in particular synaptic oligomers in human brain, which is an important knowledge gap to address to develop tau-directed therapeutics to protect synapses from degeneration. Phosphorylated and misfolded tau have previously been observed in human AD synapses using biochemical and imaging methods ^12, 16, 17^. There is a small amount of evidence from human cases suggesting that diffuse forms of tau precede NFT formation ^18^, that those diffuse forms are mainly oligomeric species ^19, 20^, and that oligomeric tau is important for seeding activity ^21^. Both high seeding activity and high concentrations of oligomers have been linked to worse cognitive outcomes ^21, 22^.

In addition to synaptotoxic effects, synaptic localisation of tau is potentially important for propagation of tau pathology through brain networks. Seminal observations of the patterns of tau pathology post-mortem by Braak and Braak ^23^ and more recent observations using tau ligands for positron emission tomography (PET) ^24^ demonstrate that NFTs accumulate in a stereotypical spatiotemporal sequence beginning in the brainstem and trans-entorhinal cortex and progressing through synaptically connected neural circuits. Since tau is secreted by presynaptic terminals in an activity-dependent manner ^25^ and misfolded tau can act as a seed to initiate pathological changes in naïve tau *in vitro* and *in vivo* ^26, 27^, a likely method of pathological propagation is via synaptic connections. It is also possible that anatomically connected brain regions are sequentially vulnerable to tau. A recent analysis of tau PET imaging and post-mortem data indicates local replication rather than spreading between brain regions may be more closely related to regional tau pathology ^28^. These two ideas are not mutually exclusive and indeed the local accumulation of tau may at least partially reflect trans-synaptic spread in local circuits, which one might predict would occur faster due to the shorter axons of local connections.

We and others have previously demonstrated trans-synaptic tau spread in model systems including transgenic mice expressing human tau in the entorhinal cortex and viral mediated tau expression ^29–33^. These studies prove that trans-synaptic tau spread is biologically possible. However, one limitation is that it might have been artificially induced by overexpression in models and this phenomenon has not been conclusively demonstrated in human brain. Here, we use sub-diffraction-limit optical imaging of synapses with multiple techniques and multiple tau antibodies (detecting oligomeric, misfolded, and phosphorylated tau) to explore in human post-mortem brain whether there is evidence of tau present in both the pre- and postsynaptic sides of the same synapses, which could be due to by trans-synaptic spread. Further we characterize tau pathology in detail to determine whether synaptic tau accumulates in areas without substantial NFTs.

## Results

### Excitatory synapses are lost in Alzheimer’s disease

Using human post-mortem brain samples prepared for array tomography (Table 1), we first replicated the observations from previous studies where synapse loss has been observed in AD ^1–3, 34, 35^. Using array tomography, we imaged 1,315,583 individual excitatory synaptic pairs in inferior temporal cortex (BA20/21). Synaptic pairs were defined as a synaptophysin-positive presynaptic puncta with a PSD95 puncta within 0.5 μm (distance between centroids). This distance was chosen based on electron microscopy data and analysis of synaptic pairs in this array tomography dataset with differing cut-off distances (Supplementary Figure 1A,B). We observed a 1.25-fold decrease in excitatory synapse density in AD when compared to controls (Control median= 4.29×10^8^±7.49×10^7^ (n=12); AD median= 3.45×10^8^±9.99×10^7^ (n=20); β=-5314.01; p=0.023; Figure 1A, B). Next, we investigated the relationship between pathological aggregates and synaptic loss in the immediate environment. To do this, regions of interest were chosen at 63x magnification with either a neurofibrillary tangle, neuritic plaque, or no aggregate in the centre of the field of view (140.6 x 106.6 μm area, (Supplementary Figure 1C). A decrease in synapse density was observed around clumps of dystrophic neurites (DNs) containing tau which surround amyloid aggregates in neuritic plaques (Control β=3862; p=0.005; AD β=3862; p=0.005; ^35, 36^). In contrast to DNs, the presence of NFTs was not associated with a decrease of excitatory synapse in the immediate environment (Figure 1A, C).

**Figure 1.**
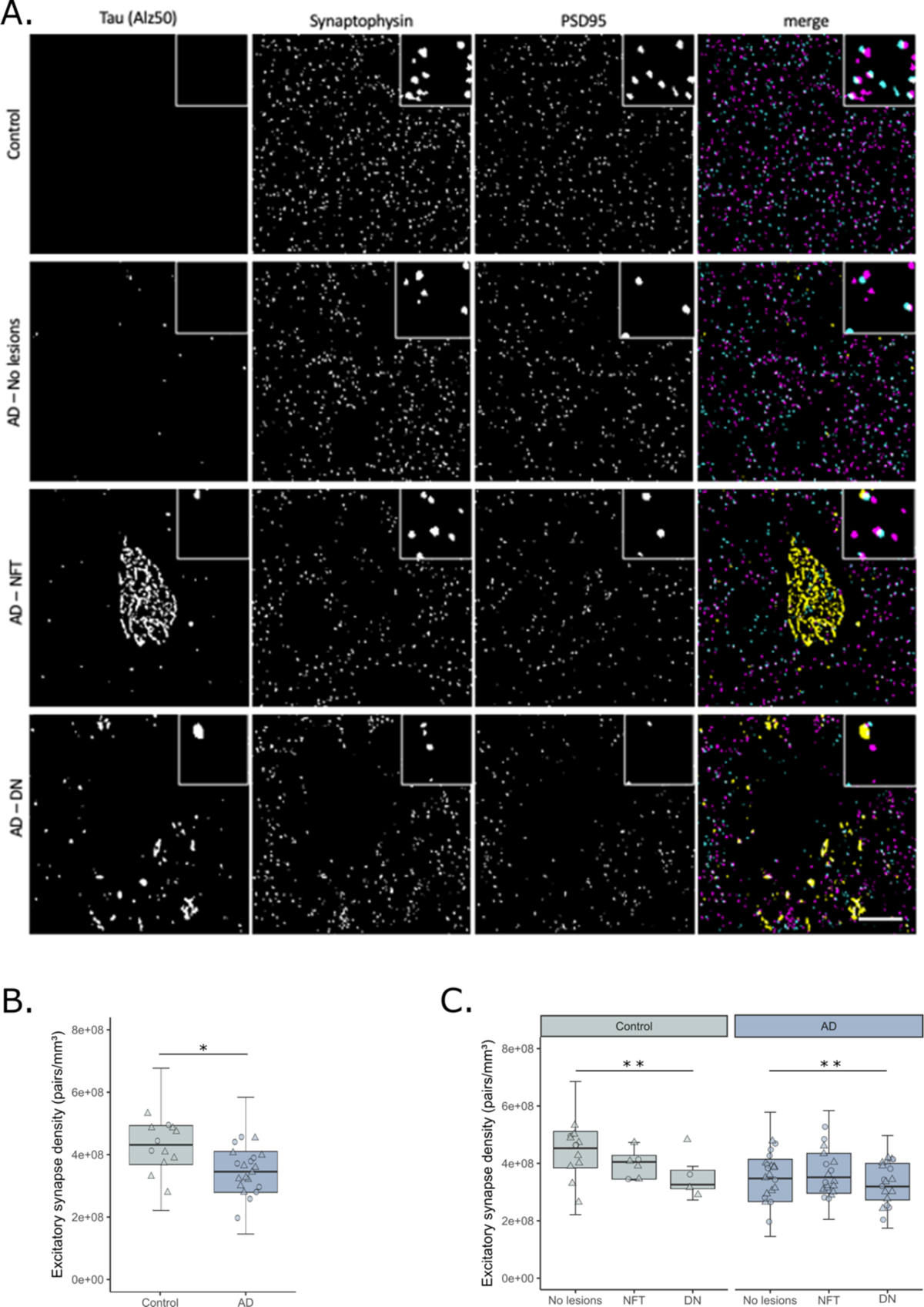
Excitatory synapse loss in BA20/21 of AD cases and its relationship to hallmark tau aggregates. **A** Representative single 70nm-thick segmented images of a control case, and AD cases with no hallmark tau aggregates, a neurofibrillary tangle or dystrophic neurite clumps around neuritic plaques. In merged images synaptophysin presynaptic terminals are shown in magenta, PSD95 postsynaptic terminals in cyan and misfolded tau (Alz50) in yellow. **B** Excitatory synapse density was decreased in AD cases when compared to controls. **C** shows the quantification of synaptic terminals split by the presence of tau aggregates. Boxplots show quartiles and medians calculated from each image stack. Data points refer to case means (females, circles; males, triangles). Analysis was with linear mixed effects models including diagnostic group, sex and pathology in the image (no effect of sex). Scale bar: 10µm. Insets 5 x 5 μm. NFT= Neurofibrillary tangle; DN=Dystrophic neurite. *p<0.05, **p<0.01.

**Table 1.**
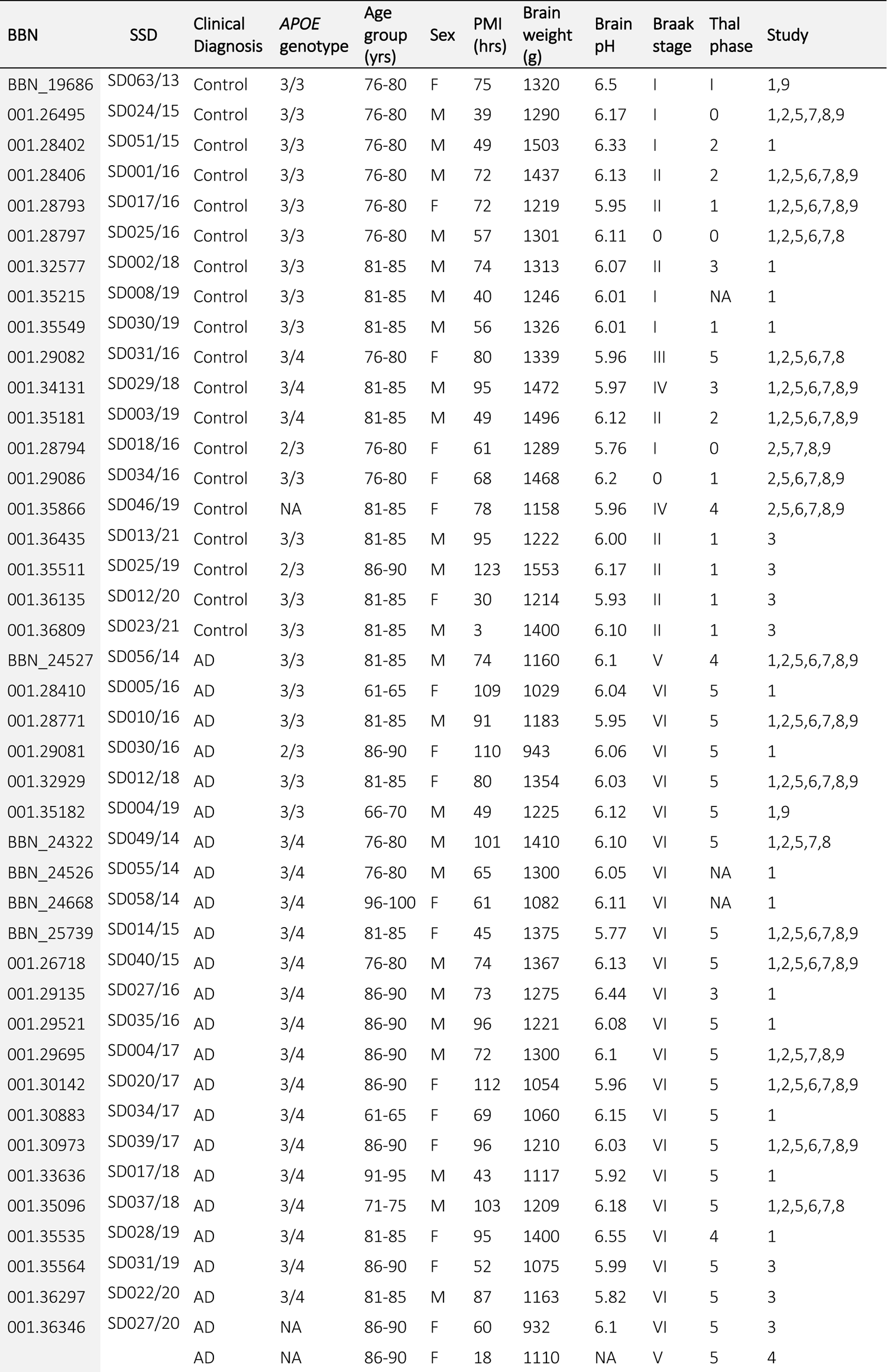
Demographic, neuropathological & genetic characteristics of human subjects AD = Alzheimer’s disease; APOE = Apolipoprotein E; BBN = Medical Research Council Brain Bank Number; SD = Sudden Death Brain Bank number; NA = not available; PMI = post-mortem interval. Study number as included in Supplementary Table 1. As part of our ethical approval with the brain banks, we are required to report anonymized case numbers where provided in the table.

### Oligomeric, misfolded, and phosphorylated tau species accumulate in postsynaptic terminals of AD cases

We next investigated the presence of tau at postsynaptic terminals. Three species of tau were studied: oligomeric (T22 antibody, characterized by ^20^), misfolded (Alz50 antibody) and Ser202, Thr205 phosphorylated (AT8 antibody) (Figure 2A-E, Supplementary figure 2A-I). All three tau species were found in postsynaptic terminals of AD cases, with a higher presence of oligomeric tau stained with T22 than the other markers (Figure 2F). Control cases also had a small amount of postsynaptic tau pathology. Interestingly, T22 and AT8 were often found in the same postsynapses while Alz50 was largely found in different synapses of AD cases (Figure 2G). There was a stronger correlation between postsynaptic tau species in AD than control cases (Figure 2G).

**Figure 2.**
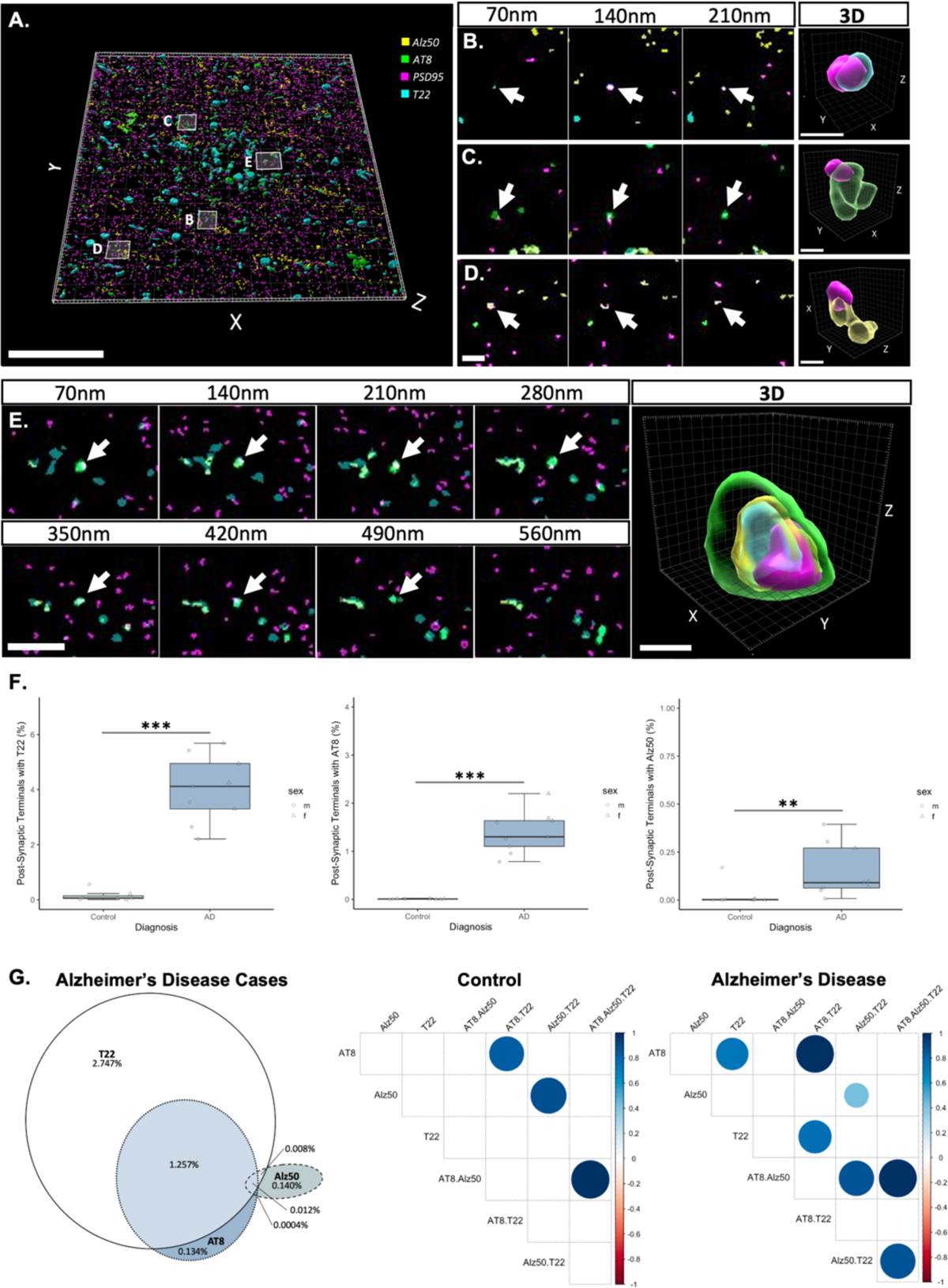
Different tau species accumulate in postsynaptic terminals. **A.** shows a three-dimensional image stack from an AD case stained with oligomeric tau (T22, cyan), misfolded tau (Alz50, yellow) and Ser202, Thr205 phosphorylated tau (AT8, green) and postsynaptic (PSD95, magenta). All three types of tau can be observed in postsynapses as shown in serial sections and three-dimensional reconstructions of individual synapses (**B-E**). Quantification (**F**) reveals increases in synaptic terminals containing tau in AD compared to control. Boxplots show quartiles and medians calculated from each image stack. Data points refer to case means (females, circles; males, triangles). \*\*\**p*<0.001, \*\**p*<0.01. A Venn diagram of the percentage of synapses that contain tau staining in AD cases (**G**, left) reveals that T22 is found in more synapses than the other species and that it colocalises often with AT8. Alz50 colocalises less with the other species. Correlation plots (**G**, right) show differences (*p*<0.01) between markers in control and AD cases. For example, synaptic T22 correlates with synaptic AT8 in AD but not in control cases.

PSD95 puncta size did not differ between control and AD cases (Supplementary figure 2J). However, there were differences in PSD95 volume when tau species were present (ANOVA after linear mixed effects model F[3,102.3]=6.85, p<0.001). All tau species had a tendency to colocalise with bigger postsynaptic terminals, with T22 being the highest observed increase, especially in control cases. We observed similar results when looking at PSD95 signal intensity as a proxy for protein levels. When looking at all postsynaptic terminals, signal intensity did not differ between control and AD cases (Supplementary figure 2K). Again, there were differences when looking at the presence of different tau species and signal intensity (ANOVA after linear mixed effects model F[3,98.3]=8.97, p<0.001). There was an overall increase of postsynaptic intensity when each tau species where present, with T22 being the highest observed increase in control cases. Taken together, both the PSD95 size and intensity seem to be related to the presence of tau species, with oligomeric tau (T22) associated with the highest increases.

The presence of tau at the synapses was confirmed with other high-resolution techniques. Immunogold electron microscopy was performed in 4 control and 3 AD cases. In all cases we observe T22-positive globular-appearing oligomers associated with fibrils in neurofibrillary tangles and on fibrils in neurites. T22 immunogold staining is also observed on presynaptic vesicles and in postsynaptic terminals near the postsynaptic density (Figure 3A, Supplementary Figure 3A). Synaptic accumulation of tau was also confirmed with dSTORM (1 control and 1 AD cases) (Figure 3B, Supplementary Figure 3D) and DNA-PAINT in an AD case (Figure 3B, Supplementary Figure 3C). Together these super-resolution imaging techniques conclusively show pathological tau in pre- and postsynaptic terminals. Finally, the presence of oligomeric tau was biochemically confirmed in synaptoneurosome preparations from AD (n=8) and control (n=8) cases (Figure 3C, Supplementary Figure 3B).

**Figure 3.**
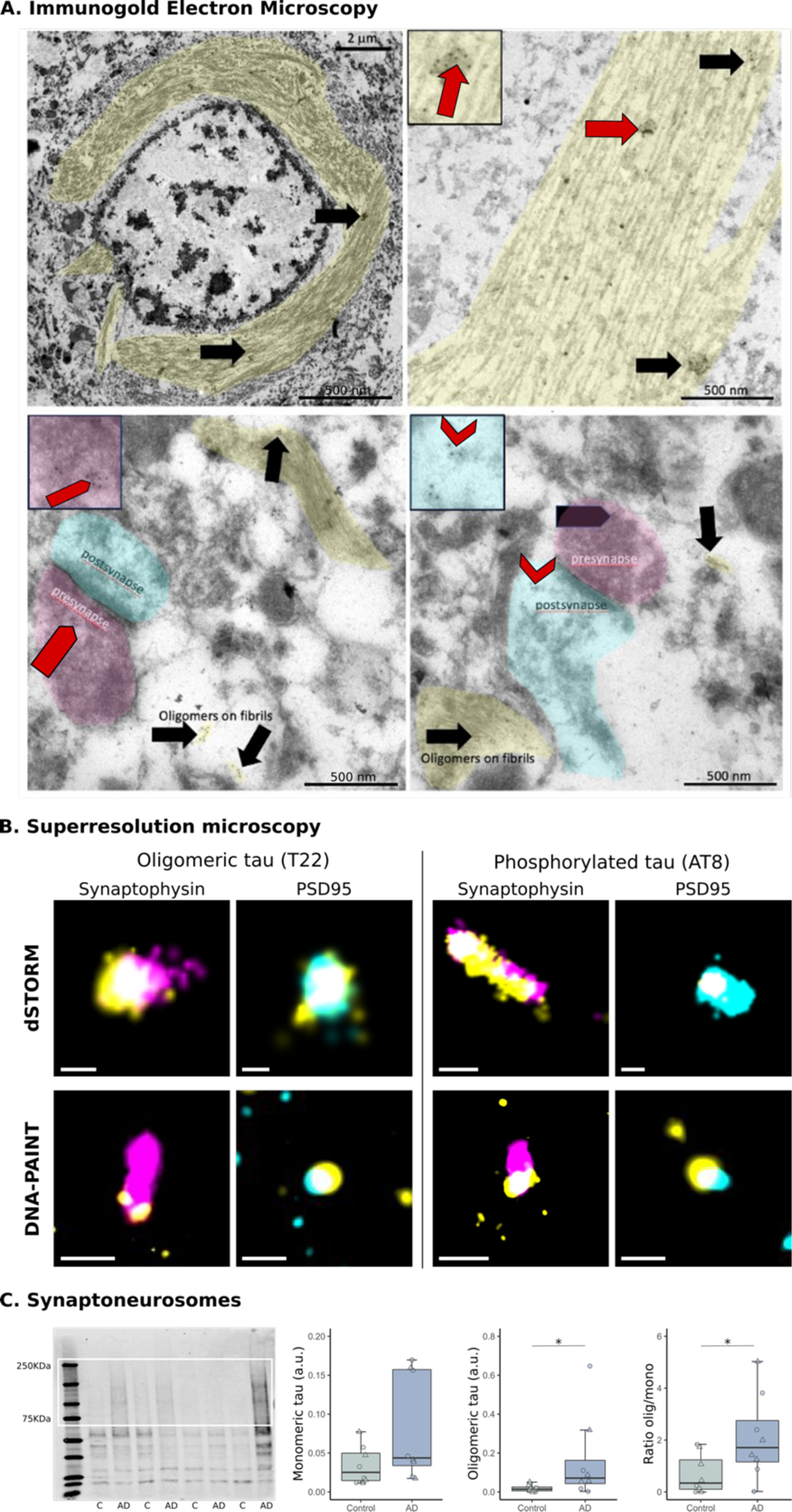
Confirmation of AD synaptic tau aggregates. **A** Immuno-electron microscopy with gold-conjugated secondary antibodies (black dots) shows T22 labels globular-appearing oligomers associated with fibrils (arrows) in neurofibrillary tangles and on fibrils in neurites (shaded yellow). T22 immunogold staining is also observed on presynaptic vesicles (pentagons, presynapses shaded magenta) and in postsynaptic terminals near the postsynaptic density (chevrons, postsynapses shaded cyan) Scale bars are labelled, insets from regions indicated with red pointers of 250nm x 250nm. **B.** dSTORM and DNA-PAINT was used on array tomography sections to confirm tau (yellow) and synaptophysin (magenta) or PSD95 (cyan) colocalization. **C.** Full western blot from synaptoneurosomes isolated from control or AD cases is shown at left and the quantification of oligomeric and/or monomeric bands at right. Boxplots show quartiles and medians (females, triangles; males, circles). *p<0.05. C = Control; SAD = Alzheimer’s disease. Scale bars dSTORM: 100nm, DNA-PAINT: 200nm.

### Synaptic oligomeric tau precedes the presence of neurofibrillary tangles

To determine whether oligomeric tau accumulation in synapses is an early pathological event, we next investigated the regional relationship between the presence of NFTs and the synaptic accumulation of oligomeric tau.

Temporal cortex (BA20/21) and primary visual cortex (BA17) samples from 10 AD cases and 10 controls were investigated for the presence of oligomeric tau at synaptic terminals by array tomography microscopy which were compared to the presence of NFTs in larger fields of view from the same brain regions by IHC on paraffin sections (Figure 4A). We found that in areas with no or low NFT counts, there was already oligomeric tau accumulating at presynaptic terminals both in control and AD cases (Figure 4A, B). There was a positive correlation between NFTs and the total amount of oligomeric tau, especially in brain areas highly affected by tau pathology (AD cases at BA20/21; *R*=0.81, *p*=0.022; Figure 4C). However, the proportion of oligomeric tau at synaptic terminals was reduced as more oligomeric tau was present, especially in BA17 where there is less overt pathology (AD cases at BA17; *R*=-0.87, *p*=0.0027; Figure 4D).

**Figure 4.**
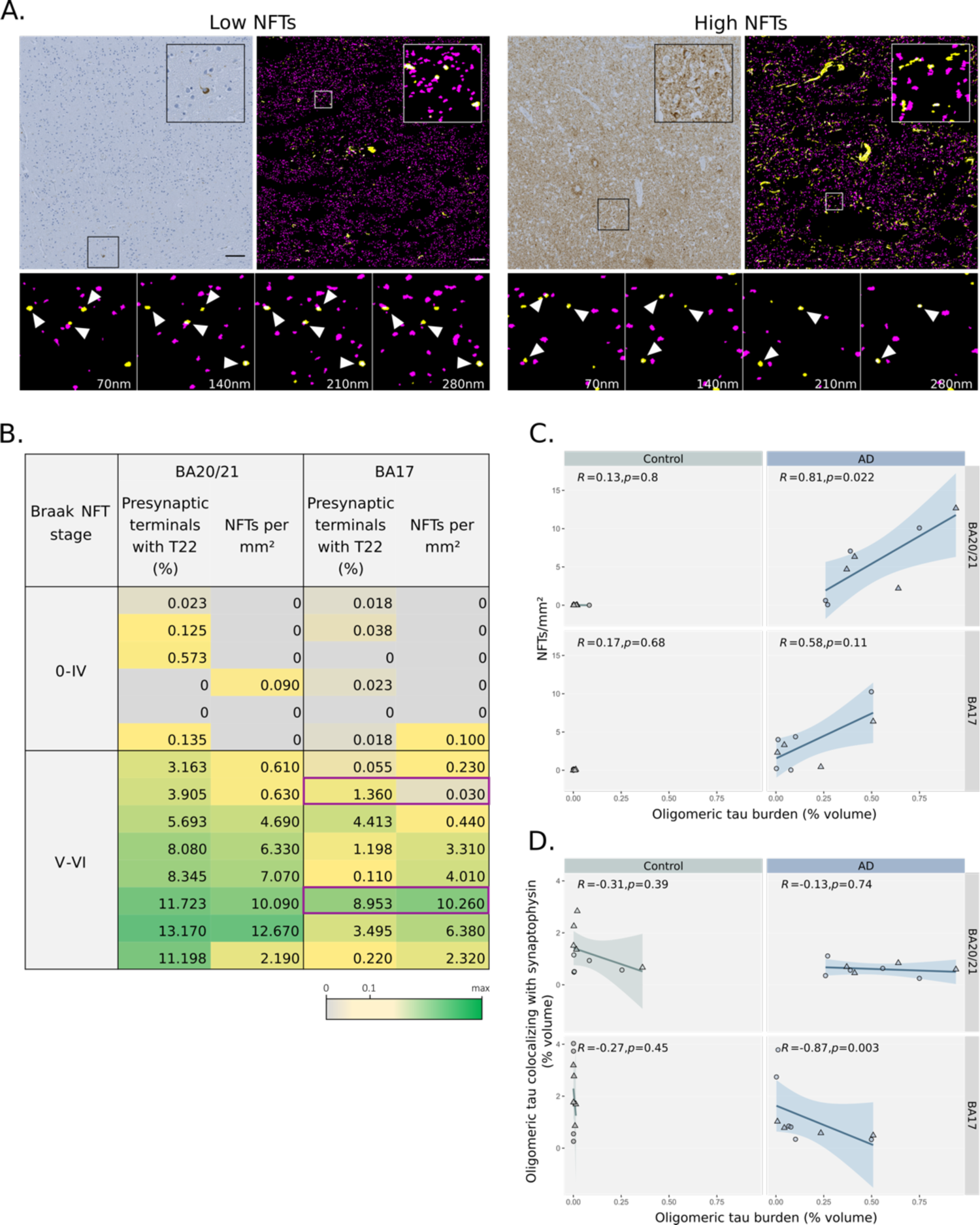
Synaptic oligomeric tau precedes the presence of neurofibrillary tangles. **A** Representative image of a case with spare (left panel) and high (right panel) NFT density. Top-left images show tau AT8 immunoreactivity by immunohistochemistry and the inset highlight the presence of NFTs. At top-right, is shown a maximum intensity projection of 12 consecutive 70nm-thick sections by array tomography microscopy. Presynaptic synaptophysin positive terminals (magenta) and oligomeric tau (T22, yellow) are shown. The inset highlights the presence of presynaptic terminals with oligomeric tau that are pointed by arrowheads in the horizontal panel of four consecutive 70nm thick sections. **B** Table displaying the density of synaptic terminals that contained oligomeric tau and the NFT density per case, area and Braak NFT stage. Highlighted in purple there are the cases shown in panel A. **C** Correlation plots between the density of NFTs and the percent volume of the image occupied by T22 and in **D** correlation plots between the percent of T22 that colocalize with presynaptic terminals and the total amount of T22. Spearman correlation results are shown in each condition. Data points refer to case means (females, circles; males, triangles). Scale bars: left 100µm, right 10µm. Insets size: left 160µm x 160µm, right 10µm x 10µm.

To confirm whether synaptic oligomeric tau is an early phenotype, we compared staining of AT180 which labels tau phosphorylated at residue 231 (reported to be an early marker of tau pathology ^37^) with oligomeric tau in synapses. While both types of tau are more prevalent in AD than control synapses, we observe 3.3 fold more T22 than AT180 labelled presynaptic terminals in BA20/21 of AD cases (presynaptic terminals with T22: Control median= 0.297±2.038% (n=9); AD median= 5.061±3.389% (n=9); β=4.15; p=0.004; Supplementary Figure 4B. Presynaptic terminals with AT180: Control median= 0.0219± 0.697% (n=9); AD median= 1.534± 1.441% (n=9); β= 1.537; p=0.01; Supplementary Figure 4C).

### Indirect evidence of transsynaptic transmission of tau aggregates

The finding of tau accumulating at synapses of AD cases even in areas with little tau pathology is in line with the hypothesis that tau may spread trans-synaptically. To further explore this possibility, the subset of tau-containing excitatory synaptic pairs were explored for pre- and postsynaptic location of tau (Figure 5A). Oligomeric tau was differentially located in the synaptic compartments (ANOVA after linear mixed effects model F[2,88]=73.61, p<0.0001); it was more commonly found in presynaptic terminals only, with a gradient towards postsynaptic localization (Figure 5B). In inferior temporal cortex, BA20/21, we confirmed that misfolded tau antibody (Alz50) was also found in both sides of the synnapse (Supplementary Figure 5A-E). Finally, to overcome the descriptive nature of post-mortem studies, we investigated transsynaptic spreading of oligomeric tau in an animal model. We previously demonstrated that human tau can spread from pre- to postsynaptic terminals in a mouse model with overexpression of human P301L mutant tau and synaptophysin tagged with GFP in entorhinal cortex^32^. Here, tissue from this same mouse model was stained with T22 to show that oligomeric tau can spread from pre- to postsynaptic terminals in mammalian brain (Supplementary Figure 5F).

**Figure 5.**
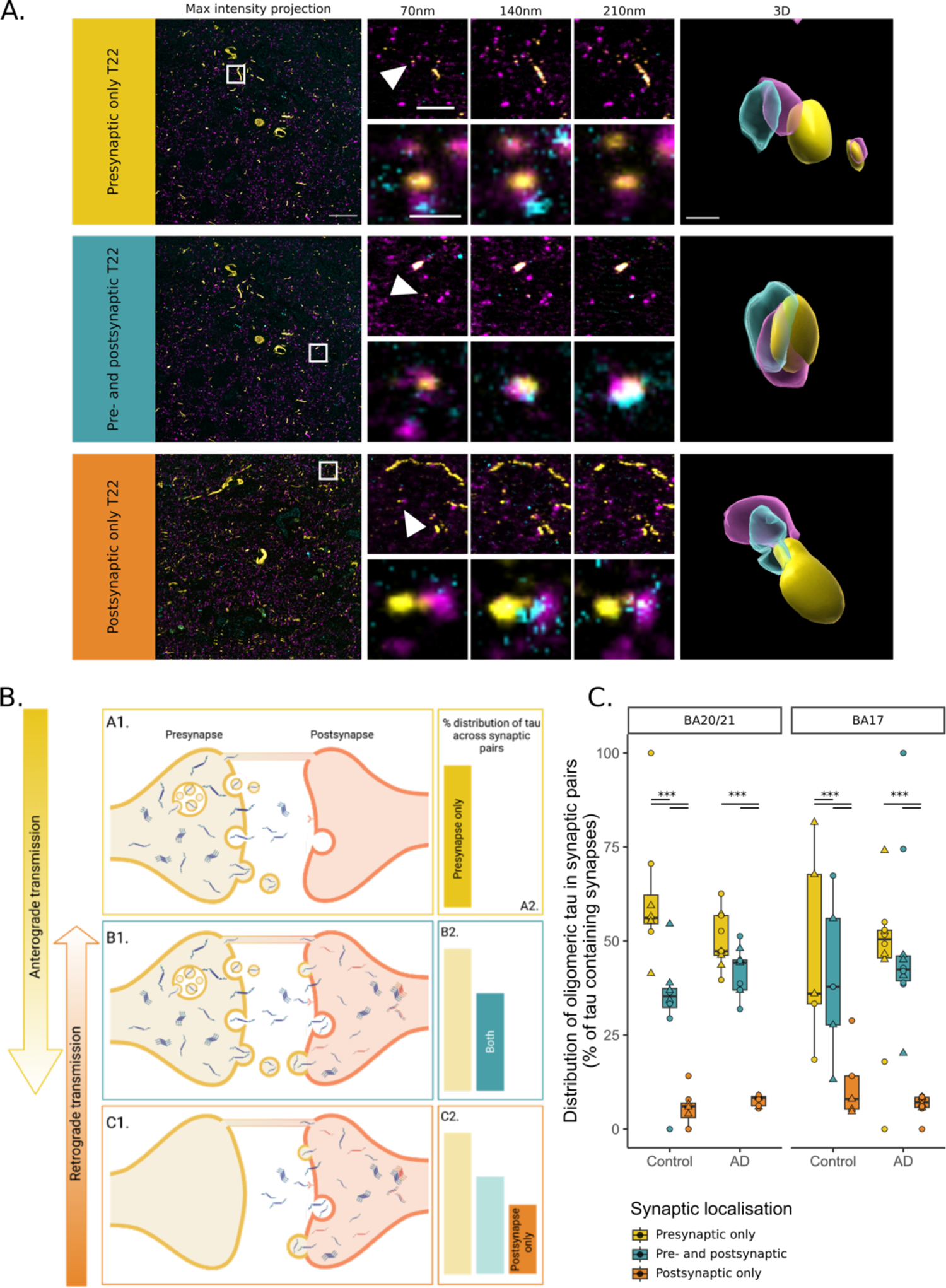
Insights into transsynaptic tau transmission. **A** Raw images from array tomography show the presence of pre (magenta) and post synaptic terminals (cyan) with oligomeric tau (yellow). First column shows a maximum intensity projection of raw images with a white box outlining the inset shown at top right of every image. Three consecutive 70nm thick sections are shown for the inset and the synapse pointed by the arrowhead is shown below and at the last column in a 3D reconstruction. **B** Diagram of the possible conditions found in tau synaptic distribution. **C** Distribution of tau from pre- to postsynaptic terminals by diagnosis and brain region. Boxplots show quartiles and medians calculated from each image stack. Data points refer to case means (females, circles; males, triangles). Analysis was with linear mixed effects models including diagnostic group, sex and pathology in the image. ***p<0.0001 in post-hoc emmeans test after linear mixed effects model. Scale bars: 20µm (raw image), 5µm (first inset), 1 µm (second inset), approximately 500 nm (3D).

## Discussion

In the present work, we show that oligomeric, phosphorylated and misfolded tau can be detected within synapses of human AD cases. As far as we know, this study represents the first comprehensive visualization of oligomeric tau at synaptic terminals in human tissue. The presence of oligomeric tau at synaptic terminals was observed even in cases with low NFT pathology and represented a higher proportion of the tau burden in regions with little NFT pathology, suggesting an early synaptic localisation of oligomeric tau. The distribution of oligomeric tau at synapses was asymmetric with the highest proportion in presynapses with a gradient towards the postsynapse. This is indirect evidence for transsynaptic anterograde transmission of tau.

### Synaptic loss and synaptic tau

Synaptic loss and alterations in synaptic markers represent one of the earliest findings in AD-affected brain regions and are the best pathological correlate of cognitive decline ^2,38, 39^. Here, we used array tomography microscopy ^40, 41^, immunogold-electron microscopy, dSTORM, and DNA paint, which allowed us to resolve different tau species in single synaptic terminals. With this approach we first replicated the synaptic loss associated with AD, as we and others have shown with several techniques ^34, 35, 39^. Previous studies using array tomography in human post-mortem brain identified synapse loss in areas surrounding amyloid beta (Aβ) plaques, with synapse loss correlating with distance from the plaque ^34, 35^. In this study, paired presynaptic and postsynaptic terminals were reduced in AD and reduced further near neuritic plaques identified by surrounding dystrophic neurites. In contrast, we did not observe synapse loss in the direct vicinity of neurofibrillary tangles. While amyloid plaques are extracellular lesions surrounded by oligomeric Aβ that is known to be toxic to synapses, neurofibrillary tangles are intracellular lesions which are thought to be harmful to the neuron containing them or to reflect a sequestering of toxic tau. This toxicity could induce loss of synapses formed by the tangle-bearing neuron, which are typically distant from the cell body, sometimes even in the opposite hemisphere. Some of the postsynaptic dendritic spines of tangle-bearing neurons are likely within the same image stack as the soma, but many of these will also be out of frame. We thus postulate that while plaques cause local synaptic toxicity, tau-induced synaptotoxicity is more diffuse throughout the brain. Second, we found that oligomeric, phosphorylated and misfolded tau all accumulate in synaptic terminals. We observed an increase in signal intensity and volume in tau-containing postsynaptic terminals, which may reflect compensation for the injured environment, something that has been suggested previously by others ^38, 42^. However, we need to be cautious interpreting these data, as light microscopy techniques are sensitive and subtle changes may affect both size and intensity. We have previously observed synaptic tau with antibodies raised to total tau, misfolded tau (Alz50) and tau phosphorylated at serine 202 and threonine 205 (AT8) ^11–13^. Here, we combined them and found that oligomeric tau is the most common tau species in postsynapses. This is an important finding as animal studies indicate oligomeric tau is particularly synaptotoxic ^15^. In previous post-mortem studies, analysis of synaptosomes ^43–47^ and synaptic fractions of homogenised tissue ^48^ have identified tau to be localised to both synaptic compartments in control and AD tissue. However, methods involving biochemical isolation of synapses fail to consider the overall architecture of the intact tissue and may unwittingly select for certain populations of synapses. Tissue homogenisation can also be damaging to synapses, allowing soluble proteins to leak in or out of the preparations. Further, this method does not generally allow for study of multiple proteins within the same synapse. Array tomography overcomes some of these limitations, offering a means for high-resolution characterisation of a large number of synapses in situ in human post-mortem tissue, and enables the assessment of synaptic density and protein composition ^40^.

The oligomeric tau we detected with immuno-electron microscopy is remarkably similar in morphology to oligomers biochemically isolated from human AD extracellular vesicles observed by atomic force microscopy ^49^. Here we demonstrate that globular structures containing oligomeric tau are associated with fibrils and within some synapses. We also observe tau labelling of synaptic vesicles consistent with observations in *Drosophila* that tau binds synaptic vesicle proteins and our previous work showing with array tomography that tau is present in presynaptic terminals ^12, 13^. This finding was further confirmed with dSTORM and DNA-PAINT that both demonstrated colocalization between pathological tau and presynaptic vesicle proteins with nanoscale resolution.

The finding of tau accumulating at synapses, and especially oligomeric tau, may have implications for the understanding of AD pathogenesis. While many studies have focused on Aβ as the trigger for AD-related synaptic damage, tau can mediate synapse loss, both alone and in concert with Aβ ^50^. Most tauopathy models show synapse dysfunction and loss ^7,51–53^. And mounting evidence suggests that oligomeric, misfolded forms of tau mediate synaptotoxicity. Tau oligomers may induce morphological changes, impair plasticity and dysregulate synaptic transmission ^54–57^. Oligomers also induce synapse loss and impairment in memory function, which is rescued upon reduction of oligomer levels ^58–60^. Thus, at the synapse, tau oligomers may reduce synaptic proteins, alter neuronal signalling, promote synapse loss and impair memory function.

In this study, we also found oligomeric tau in cases with no or low NFT presence. These findings are in line with evidence from human cases suggesting that diffuse and neuropil forms of tau precede NFT formation ^18, 37^ and may involve oligomeric species ^19, 20^. This early (according to NFT burden and the well stablished Braak staging) oligomeric tau was found to accumulate at synapses. The seeding activity of tau in synaptoneurosome preparations has also been found to precede the amount of hallmark pathology as reflected by Braak stages ^61^ and seeding closely correlated with oligomeric tau ^21^. Of note, both high tau seeding and high levels of tau oligomers have been linked to worse cognitive outcomes ^21, 22^. We also found that the proportion of oligomeric tau in presynaptic terminals was inversely correlated with the total burden of oligomeric tau. Paralleling Braak staging, these findings may imply that i.e. while primary visual cortex does not have enough pathology to be classified as Braak stage VI, we already find oligomeric tau at synapses that may be affecting neuronal circuits.

Synaptic localisation of tau may be also important for propagation of tau through the AD brain. Oligomeric tau was distributed asymmetrically across synaptic pairs. This possibly suggests a model where tau accumulates presynaptically, before release and uptake at the postsynapse, leading to tau localisation in both compartments. It is unlikely that upon transference to the postsynapse, pathological tau would be completely lost from the presynaptic terminal. Thus, synaptic pairs in which tau is localised to the postsynapse only could arise from retrograde transport to the postsynapse (i.e., along dendrites and spines) ^62^. Further, tau could undergo retrograde transport across the synapse, to the presynaptic site ^63, 64^. However, considering the stepwise decrease in oligomeric tau localisation observed at synaptic pairs (presynapse only > both terminals > postsynapse only), it is likely that tau predominantly traverses synapses in the anterograde direction. Oligomeric tau followed this pattern of distribution across synaptic pairs in both control and AD brain. However, the proportion of synapses with tau was significantly increased in AD brain, indicating that although the mode of spread may be the same, propagation likely occurs at greater frequency in AD. While these findings gave us insights into the pathogenesis of AD and can have implications in therapeutical approaches, we need to be cautious due the inherent descriptive cross-sectional nature of post-mortem studies.

## Conclusions

In the present work we found that oligomeric, misfolded, and phosphorylated tau species are found at synaptic terminals of AD cases even when NFT burden is low. Given the potential early nature of these events in AD pathogenesis, we believe that these findings support the notion of multiple target and synaptic based approaches for therapeutic interventions.

## Data Availability

All data produced in the present study are available upon reasonable request to the authors or available online at the University of Edinburgh DataShare repository.

https://github.com/Spires-Jones-Lab

https://datashare.ed.ac.uk/handle/10283/3076

## Acknowledgements

We gratefully acknowledge the contributions of our brain tissue donors and their families, the Edinburgh Brain and Tissue Bank and Alzheimer’s Scotland Dementia Research Centre for coordinating brain tissue donations, and Prof Rakez Kayed for generously providing T22 antibody. This work was supported by the Medical Research Council Centres of Excellence in Neurodegeneration (CoEN5025), the European Research Council (ERC) under the European Union’s Horizon 2020 research and innovation programme (grant agreement no. 681181) and the UK Dementia Research Institute which receives its funding from DRI Ltd, funded by the UK Medical Research Council, Alzheimer’s Society, and Alzheimer’s Research UK. The confocal microscope was generously funded by Alzheimer’s Research UK and a Wellcome Trust Institutional Stategic Support Fund at the University of Edinburgh. JL was funded by UCB Biopharma, as was the Oxford Nanoimager.

## STAR Methods

### Resource availability

#### Lead contact

Further information and requests for resources and reagents should be directed to and will be fulfilled by the lead contact, Prof. Tara Spires-Jones.

#### Materials availability

This study did not generate new unique reagents.

#### Data and code availability

- Raw images are available from the corresponding author upon reasonable request. Original western blot images are included in the supplementary data.
- Protocols, image analysis scripts and R scripts for statistical analysis are freely available (https://github.com/Spires-Jones-Lab).
- Any additional information required to reanalyze the data reported in this paper is available from the lead contact upon request.

### Experimental model and subject details

#### Human brain tissue

Brain samples were obtained with ethical approval via the University of Edinburgh Sudden Death Brain Bank and the Alzheimer Scotland Brain and Tissue Bank. This study was reviewed and approved by the Edinburgh Brain Bank ethics committee and the Academic and Clinical Central Office for Research and Development, a joint office of the University of Edinburgh and NHS Lothian (approval number 15-HV-016). The Edinburgh Brain Bank is a Medical Research Council funded facility with research ethics committee (REC) approval (16/ES/0084). As part of our ethical approval with the brain banks, we are required to report anonymized case IDs as in Table 1.

Inclusion criteria for Alzheimer’s disease cases were a clinical dementia diagnosis, Braak stage V-VI, and neuropathological diagnosis of AD. Control subjects were donors without neurological or psychiatric diagnoses and were included based on age, sex, and *APOE* genotype matching the AD cases as closely as possible. All cases were genotyped in-house to determine *APOE* genotype, as described previously ^65^. Exclusion criteria for both groups were neuropathological findings indicative of substantial secondary pathology (e.g., haemorrhage or Lewy bodies within brain region of interest). A total of 23 AD and 19 control cases were examined in this study as outlined in Table 1 showing demographic data and the experiments they were used in. Sex was included as a fixed effect in statistical models to examine sex effects in all measures.

A statistical power analysis was performed for sample size estimation, based on data from study 1 (Table 1, Supplementary Table 1), comparing presynaptic terminals with tau to diagnosis (control or AD) by linear mixed effect model (effect size *β*=0.521, see statistical analysis section). With an alpha= 0.05 and power = 0.80, the projected sample size needed with this effect size in approximately N= 10. The analysis was performed as described in ^66^.

Brain samples from inferior temporal cortex (Broadmann Areas 20 and 21, BA20/21) and primary visual cortex (BA17) were collected at autopsy and preserved by fixation and paraffin embedding, and fixation for array tomography as detailed previously ^67^. These regions were selected to compare areas with high pathological burden (BA20/21) to those with low pathology (BA17).

#### Mouse brain tissue

Mice expressing human P301L mutant tau, cytoplasmic tdTomato, and green fluorescent protein tagged synaptophysin in entorhinal cortex neurons were used to determine whether oligomeric tau can spread anterogradely from pre to post synapses. Tissue was used from mice generated in a previous study ^32, 68^. Briefly, FVB-Tg(tetO-TauP301L)4510 mice ^69, 70^ were crossed with the Tg(tetO-tdTomato-Syp/mut4EGFP)1.1Luo/J line from Jackson laboratories. Offspring expressing both Tg(tetO-TauP301L)4510 and Tg(tetO-tdTomato-Syp/mut4EGFP) transgenes were crossed with a line expressing the tetracycline sensitive transcriptional activator controlled by the *Klk8* neuropsin promotor (EC-tTA), which expresses the tTA heavily in the EC and not at all in the dentate gyrus ^71^. These crosses generated mice expressing both rTgTauEC+EC-tdTomato/Syp-GFP. Blocks containing entorhinal cortex and hippocampus were sectioned for array tomography from both male and female animals at 3 months (n=3), 9 months (n=4) and 18 months (n=4). A mouse without P301L Tau expression which did express tdTomato and GFP was included as a negative control for T22 staining and there was a no primary negative control condition. Animals were maintained on a 12h light/dark cycle with ad libitum access to food and water. All animal experiments were approved by the Harvard Medical School Institutional Animal Care and Use Committee and the UK Home Office.

### Method details

#### Array tomography

Tissue samples were fixed in 4% paraformaldehyde for 3 hours, dehydrated, and embedded in LR White resin. 70nm serial sections were cut with a diamond knife (Diatome) using an ultracut microtome (Leica). Ribbons of 15-30 serial sections were collected onto gelatin coated coverslips and immunostained with antibody combinations shown in Supplementary Table 1. Images were obtained with a 63x 1.4 NA objective on an AxioImager (Zeiss) or Leica TCS confocal (Leica). In study 1, images were taken in regions containing NFTs, dystrophic neurites surrounding plaques, or no pathological lesions. Pathological lesions were identified by 2 experienced investigators (CD and MCC) based on international guidelines as we have used previously (Supplementary figure 1) ^72–74^. Only one microscope was used for imaging each experimental condition. Image from the same location in each serial section along a ribbon were aligned, thresholded, and parameters quantified using in house scripts in Image J, MatLab (Mathworks), and Python. All software is freely available on GitHub (https://github.com/Spires-Jones-Lab). Saturation was minimized during image acquisition and only applied for figure visualization. 3D reconstructions were performed with Imaris software (Bitplane).

#### Immunogold-electron microscopy

Tissue samples were collected at autopsy and fixed in 4% paraformaldehyde and 0.1% glutaraldehyde in 0.1M phosphate buffer (PB) for 48 hours at 4°C. After fixation, the tissue was washed with 0.1M PB twice and stored at 4°C prior to processing and embedding. Tissue processing was carried out using an Electron Microscope Tissue Processor (Leica). In the processor, the tissue was first placed in 0.1% osmium tetroxide (O014, TAAB Laboratories) in boiled distilled water for 1 hour. Thereafter, samples were dehydrated in a series of solutions in preparation for embedding: 3 cycles of 5 minutes each in boiled distilled water, 50% ethanol, 70% ethanol and 90% ethanol. This is followed by 2 cycles of 15 minutes in 100% ethanol, and 2 cycles of 2 minutes in propylene oxide. The samples were then placed in a mixture of half propylene oxide (AGR1080, Agar Scientific) and half LR white resin (AGR1281, Agar Scientific) for 30 minutes, before being transferred to a full solution of LR white resin for 2 cycles of 30 minutes. These tissues were then taken out of the processor, placed in a gelatin capsule (C087/1, TAAB Laboratories), and filled up with LR white resin to the brim. Polymerisation of the LR white resin was allowed to occur by thermal curing at 52°C for 24 hours. These resin-embedded tissue blocks were then stored at room temperature until ready for immuno-EM staining.

The resin-embedded tissue blocks were cut into 50nm ultrathin sections using an ultramicrotome (UC6, Leica) fitted with a Histo Jumbo Diamond Knife (Diatome Knives) and placed onto formvar-coated nickel grids (S162N1, Agar Scientific). Prior to immuno-EM staining, residual osmium tetroxide in the tissue were removed by incubating the tissue-covered nickel grids in saturated sodium metaperiodate in boiled distilled water for 1 minute. The nickel grids were then incubated in 1% sodium borohydride in 0.1M PB for 5 minutes to reduce free aldehydes, and thereafter incubated in 50mM glycine in 0.1M PB for 10 minutes to inactivate residual aldehyde groups. The nickel grids were then blocked in blocking buffer (0.1% BSA, 0.1% fish skin gelatin, 0.05% Tween in 0.2M PB, pH7.4) for 1 hour at room temperature. Rabbit T22 primary antibody (1:50, courtesy Dr Rakez Kayed) was diluted in antibody dilution buffer (0.1% BSA, 150mM sodium chloride in 0.2M PB, pH7.4) and incubated with the nickel grids overnight at 4°C. The next day, the nickel grids were washed in 6 rinses of 0.2M PB. Goat anti-rabbit 10nm gold-conjugated secondary antibody (1:50, ab39601, Abcam) was diluted in antibody dilution buffer and incubated with the nickel grids for 90 minutes at room temperature. The nickel grids were then washed in 6 rinses of 0.2M PB, fixed in 2.5% glutaraldehyde in 0.2M PB for 15 minutes, and washed in 6 rinses of 0.2M PB again. Negative staining of the nickel grids was carried out to improve imaging contrast. This was done in a carbon dioxide free environment: incubated the nickel grids with 3% uranyl acetate in 50% ethanol for 15 minutes and 3% lead citrate in boiled distilled water for 150 seconds, rinsed thrice in boiled distilled water in between the steps. Once the nickel grids were dry, images were captured using a Transmission Electron Microscope (JEM-1400 Plus, JOEL) and viewed using ImageJ (Version 1.44).

#### Direct STORM acquisition and image reconstruction

For the reconstruction of the images using direct STORM, sections 70 nm thick were stained as previously described ^75^, using the primary antibodies T22 (Millipore) or AT8 (Innogenetics) and Synaptophysin (R&D Systems), and the secondary antibodies Alexa 488 (ThermoFisher) and Alexa 647 (ThermoFisher), finally stained with Hoechst 33258 (Life Technologies) to visualise the nuclei, then the coverslips were mounted with OXEA buffer ^76^. STORM images were acquired using a Nikon N-STORM system configured for total internal reflection fluorescence (TIRF) imaging. Laser inclination was tuned to adjust focus and to maximize the signal-to-noise ratio. AlexaFluor-647 was excited illuminating the sample with the 647 nm (20% of 160 mW) and AlexaFluor-488 with the 488 (40% of 80 mW) lasers line built into the microscope. Fluorescence was collected by means of a Nikon 100x, 1.49 NA oil immersion objective and passed through a quad-band pass dichroic filter (97335 Nikon). 10,000 frames at 20 ms integration time were acquired for each channel. Images were recorded onto a 256 × 256 pixel region of a Hamamatsu ORCA Flash 4.0 CMOS camera (0.16 μm pixel size). STORM images were analysed with the STORM module of the NIS element Nikon software ^77^.

#### DNA-PAINT

DNA Conjugation to the AT8 and T22 antibodies and donkey anti-rabbit IgG (H+L) cross-absorbed secondary antibody (Invitrogen, 31238) was performed using the Siteclick Antibody Azido modification Kit (Thermo Fisher Scientific, S20026) according to the manufacturer’s instructions. Briefly, after buffer exchange, 200 µg of the AT8, T22 antibodies and secondary antibody were incubated for 6 h at 37 °C with ß-galactosidase. To conjugate an azido group to the antibodies, they were incubated over-night at 30 °C in a mixture of UDP-GalNAz and GalT enzyme. After purification, the modified antibodies were incubated with DBCO-DNA (2-ACCACCACCACCACCACCA, 2 = 5’ DBCO TEG, ATDBio Ltd.) in a 1:5 ratio over-night at 25 °C. The remaining unconjugated DNA was removed with 100 kDa Amicon columns (Merck, UFC510024).

Frame seal slide chambers (9 × 9 mm, Biorad, Hercules, USA) were affixed to the AT sections onto the coverslip, which were stained as previously described (Kay K. R. et al, 2013a) 1) using AF488-labelled synaptophysin (1/100 dilution) and either DNA-conjugated AT8 (100 nM) or DNA-conjugated T22 (100 nM) antibodies or 2) using PSD95 (Cell Signaling Technology, D27E11 XP rabbit mAb) (1/50 dilution) and DNA-conjugated donkey anti-rabbit IgG (H+L) cross-absorbed secondary antibody with either DNA-conjugated AT8 (100M) or DNA-conjugated T22 (100nM) antibodies. ATTO655-tagged imager strand (GGTGGT-ATTO655, ATDBio Ltd.) was diluted in GLOX buffer (40 μg/mL Catalase, 0.5mg/mL glucose oxidase, 10% w/v glucose and 200 mM β-mercaptoethanol) and used at a final imaging concentration of 2nM or 4 nM. Cy3B-tagged imager strand (AGAGAGAX-Cy3B, ATDBio Ltd.) was diluted in imaging buffer (5 mM Tris-HCl pH 8.0, 1mM EDTA, 75 mM MgCl2, 0.05% Tween-20) and at a final imaging concentration of 2 nM.

DNA-PAINT for synaptophysin and AT8 or T22 combined with dSTORM was performed on an Oxford NanoImager (ONI) super-resolution microscope equipped with four laser lines (405, 488, 561, and 638 nm) and a 100× oil-immersion objective (Olympus 1.49 NA). Fluorescence images were acquired by exciting multiple 50 µm × 80 µm areas with 488 nm laser for AF488-labelled synaptophysin (dSTORM, laser power 44–47 mW, interval excitation with 405 nm laser per 200 frames, 5,000 frames with an exposure time of 50 ms) or 638 nm laser for ATTO655-tagged imaging strands targeting the conjugated AT8 or T22 antibodies (DNA-PAINT, laser power 60–66mW, 5,000 frames with an exposure time of 50 ms). The laser was set at 53° incidence angle. Images were recorded by NimOS software associated with the ONI instrument.

DNA-PAINT for PSD95 and AT8 or T22 was performed using a custom-built TIRF microscope, restricting excitation of fluorophores within the sample to 200 nm from the sample-coverslip interface. The fluorophores were excited at either 561 nm (Cy3B) or 638 nm (ATTO655). Collimated laser light at wavelengths of 561 nm (Cobolt DPL561-100 DPSS Laser System, Cobalt, Sweden) and 638 nm (Cobolt DPL638-100 DPSS Laser System, Cobalt, Sweden) were aligned and directed parallel to the optical axis at the edge of a 1.49 NA TIRF Objective (CFI Apochromat TIRF 60XC Oil, Nikon, Japan), mounted on an inverted Nikon TI2 microscope (Nikon, Japan). A perfect-focus system corrected the imaging process for any stage-drift. Fluorescence was collected by the same objective and separated from the TIR beam by a dichroic mirror Di01-R405/488/561/635 (Semrock, Rochester, NY, USA). Collected light was then passed through appropriate filters (561 nm: LP02-568-RS, FF01-587/35 (Semrock, NY, USA, 638 nm: FF01-432/515/595/730-25, LP02-647RU-25 (Semrock, NY, USA)). The emission beam was passed through a 2.5x beam expander and focussed onto an EMCCD camera for image collection (Delta Evolve 512, Photometrics, Tucson, AZ, USA) operating in frame transfer mode (EMGain= 11.5 e^-^/ADU and 250 ADU/photon). Pixel size was 103 nm. Images were recorded with an exposure time of 50 ms with 638 nm illumination, followed by 561 nm excitation. The microscope was automated using the open-source microscopy platform Micromanager (NIH, Bethesda).

#### DNA-PAINT analysis

The positions of the transiently immobilized DNA imager strands within each frame were determined using the PeakFit plugin (an imageJ/Fiji plugin of the GDSC Single Molecule Light Microscopy package (http://www.sussex.ac.uk/gdsc/intranet/microscopy/imagej/gdsc_plugins) for imageJ using a ‘signal strength’ threshold of 50 and a precision threshold of 20 nm.

#### Immunohistochemistry

Paraffin sections were stained with AT8 using the Novolink polymer detection system and visualized using 3,3’-diaminobenzideine chromogen then counterstained with haematoxylin to stain nuclei. ROIs containing cortical grey matter were delineated using QuPath ^78^ and colour deconvolved in ImageJ. Using a custom Matlab script, neurofibrillary tangles were identified minimizing the bias from manual counting.

### Synaptoneursomes isolation and Western blot analysis

Brain homogenates (H) and synaptoneurosomes (S) were prepared as described in ^46^. In brief, ∼ 200 mg of tissue from BA20/21 was homogenized on ice in 1ml homogenization buffer (25 mM HEPES pH 7.5, 120 mM NaCl, 5 mM KCl, 1 mM MgCl2, 2 mM CaCl2, protease inhibitors (Roche complete mini), phosphatase inhibitors (Millipore, 524,629). The homogenate was filtered through an 80 μm nylon filter (Millipore, NY8002500) and 300 µl was saved as crude homogenate with buffer (100mM/L Tris-HC1 pH 7.6, 4% SDS, protease inhibitor cocktail EDTA-free 100x Thermo Fisher Scientific, Loughborough, UK). The remaining crude homogenate was further filtered through a 5 μm filter (Millipore, SLSV025NB) and centrifuged at 1000 g for 5 min. The supernatant was discarded and the pellet was washed with buffer and centrifuged again, yielding the synaptoneurosome pellet. For storage it was mixed with buffer (100mM/L Tris-HC1 pH 7.6, 4% SDS, protease inhibitor cocktail EDTA-free 100x Thermo Fisher Scientific, Loughborough, UK). Protein concentrations were determined using a protein assay (Micro BCA TM Protein Assay Kit, Thermo Fisher Scientific, 23235).

Synaptoneurosomes preparations (2.5-10 μg/μl) were denatured at 95° C for 5 min. To detect oligomeric tau, 75 μg of protein were resolved using 4%–12% Bis–Tris gels (MOPS running) and transferred onto nitrocellulose membranes (Biorad, 1620112) in Trans-Blot turbo buffer (Biorad, 1704270) supplemented with 20% ethanol at 25 mV for 13 min in the Trans-Blot® Turbo™ Transfer System (Biorad, 1704150). After blocking 1h with Intercept® Blocking Buffer (Li-COR; 927-60001), the membrane was incubated with the indicated primary antibodies (α-tau5 antibody (Biolegend, 806401, 1:250) and α-GAPDH antibody (Cell Signaling, 2118,1:1000) overnight at 4°C. The protein bands were visualized using corresponding fluorescent secondary antibodies (Odyssey, 1:5,000). Infrared fluorescence was measured with the Odyssey CLx imager (LI-COR) and quantified using Image Studio software (Li-COR Biosciences).

### Study design

The immunostaining, image acquisition, image processing and analyses were performed blinded to the clinicopathological diagnosis. Bias was also minimized by setting the same parameters for image acquisition and image analysis for all cases.

#### Quantification and statistical analysis

Differences in age and PMI were assessed using Kruskal-Wallis and one-way ANOVA, respectively. Distributions of data were examined using histograms and Shapiro-Wilks tests. The majority of statistical analyses used mixed effect models (*lmerTest* package in R) to investigate the effect of disease status, *APOE* and sex on the variable of interest, while controlling for potentially confounding variables and specifying the random effect to account for repeated measures. Models were generated and residuals assessed. If data failed to meet model assumptions data was transformed using Tukey or Box-Cox transformations.

After running the mixed effects model, an ANOVA (*stats* package - Type III Analysis of Variance Table with Satterthwaite’s method) was performed on the model output to determine the main and interaction effects. If the interaction was significant or if the main effect was significant and had more than 2 levels, the *emmeans* package was used to compute multiple comparisons, where the Kenward-Rogers method for degrees of freedom and the Tukey method for P value adjustment were employed. Correlations used Pearson’s of Spearman’s methods, dependent on whether data met assumptions for parametric testing or not.

Statistical significance was set at 5% (α = 0.05). Statistical details can be found in the figure legends and results text, including the statistical tests used, exact value of n, what n represents, and dispersion and precision measures. All the analyses were performed with R ^79^ and the scripts and full statistical results can be found at \*\*\**add doi to final analysis files upon acceptance for publication*.

## Supplementary material

**Supplementary Figure 1.**
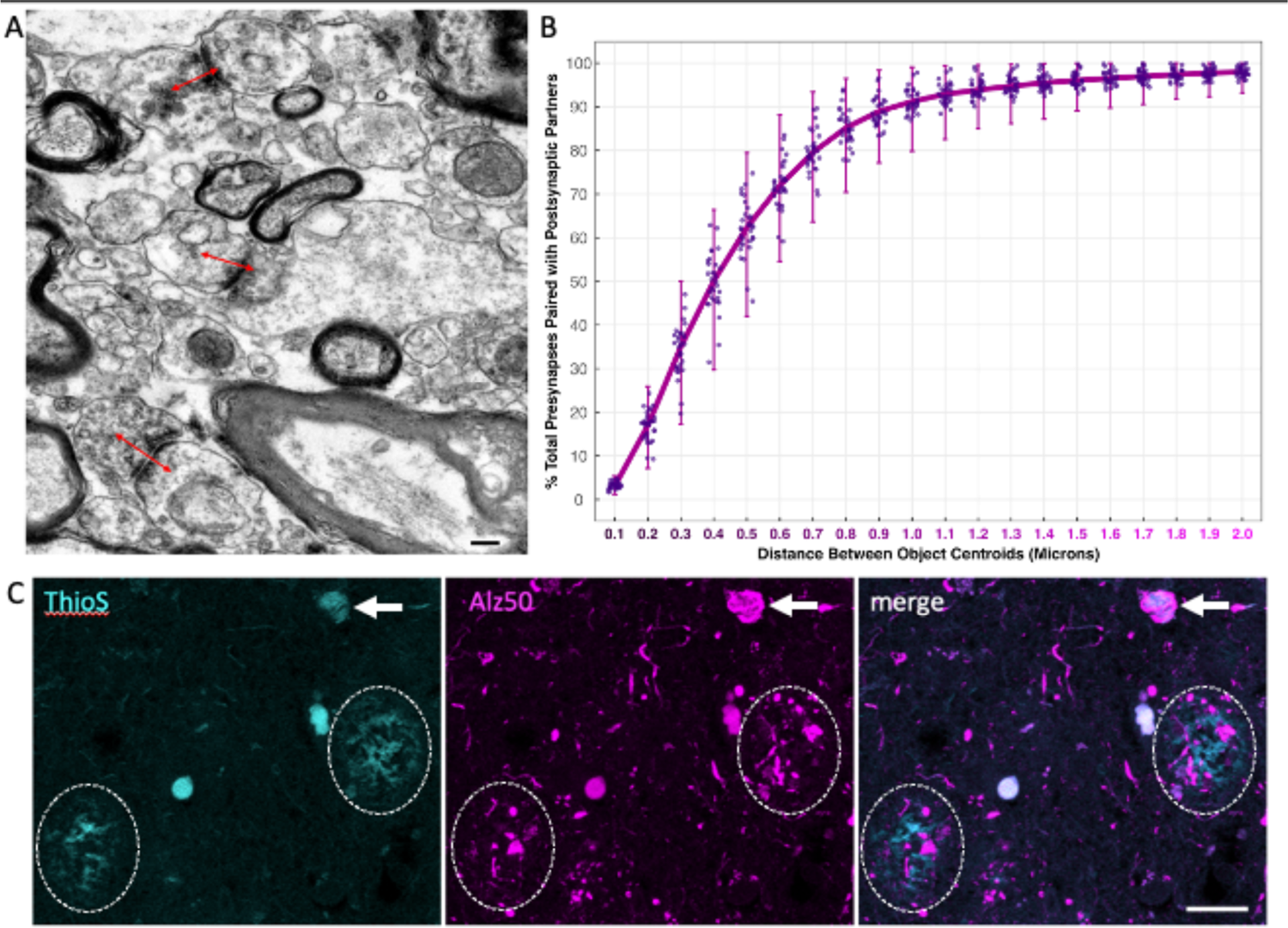
Synaptic pairs in array tomography were defined as pre and post-synaptic puncta with centroids within 500 nm (0.5 μm). This is based on electron microscopy data (example in panel **A**) showing that the centre of presynaptic terminal staining is within 500nm of the PSD of the majority of synapses (red arrows 500nm in length). Further this cut-off in our array tomography data results in 60% of presynaptic terminals having a postsynaptic partner (**B**). This is lower than the expected 80-90% excitatory synapses but reduces the chances of counting false positives which is important for inferring trans-synaptic tau spread in this study. In the array tomography study, synapse density was assessed in proximity to Alz50 positive neurofibrillary tangle pathology or neuritic plaques defined by Alz50 positive dystrophic neurite patterns. To confirm these patterns of dystrophic neurites are indeed neuritic plaques, we used confocal imaging of paraffin sections stained with thioflavin S to label plaque and tangle fibrils and Alz50 immunohistochemistry and confirmed that Alz50 labels tangles (**C**, arrows) and that Alz50 positive dystrophic neurite surround ThioS positive plaques (**C**, circles). Scale bars represent 500nm in A, 20 μm in **C**.

**Supplementary Figure 2.**
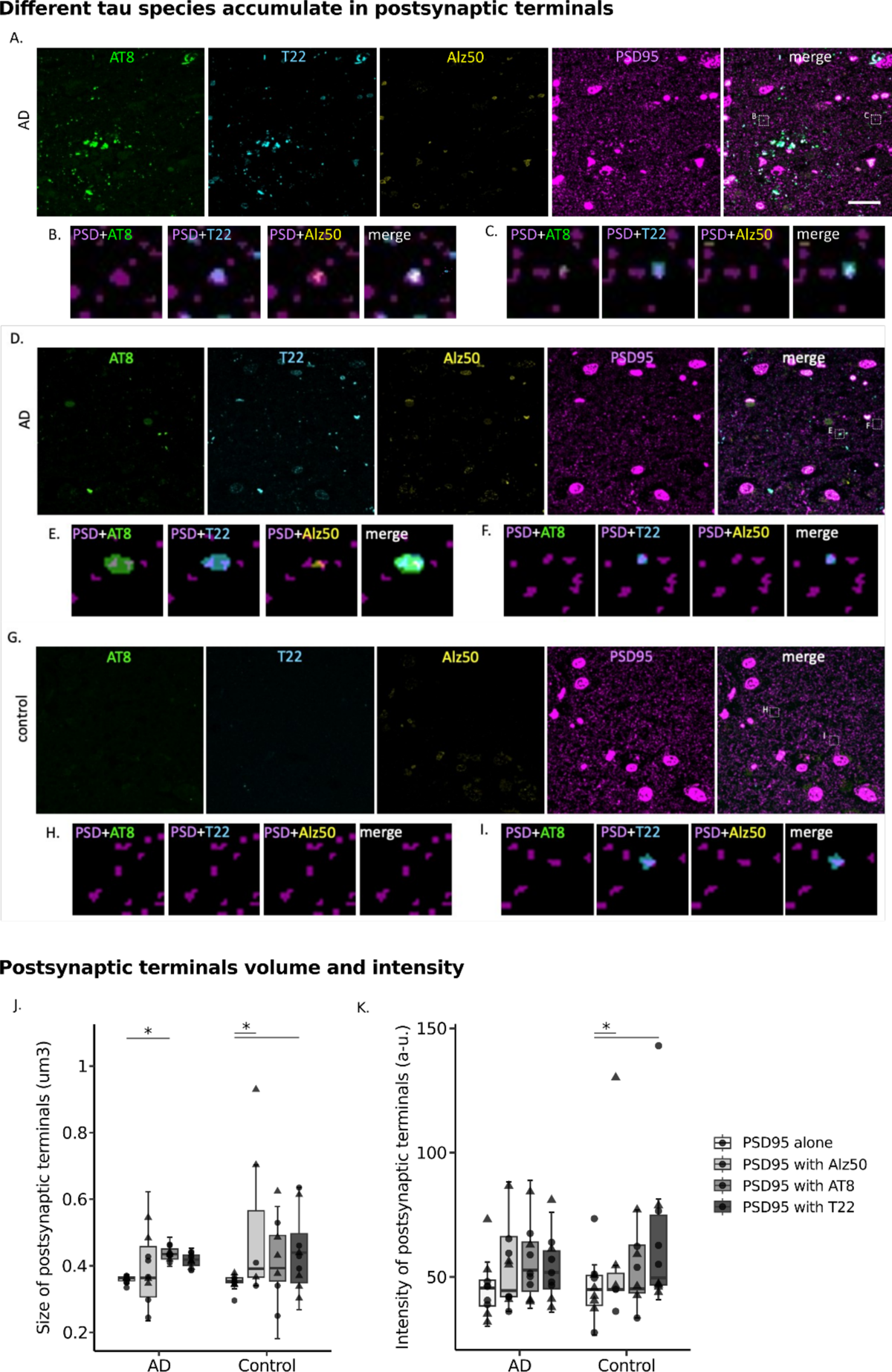
Different tau species accumulate in postsynaptic terminals. Top panel shows three more examples of postsynaptic terminals containing tau **A., D** and **G** shows a single 70nm-thick section from two AD and a control case stained with oligomeric tau (T22, cyan), misfolded tau (Alz50, yellow) and Ser202, Thr205 phosphorylated tau (AT8, green) and postsynaptic (PSD95, magenta). All three types of tau can be observed in postsynapses as shown in 5umx5um insets below (**B, C, E, F, H, I**). **Bottom panel** shows quantification of postsynaptic terminals volume and intensity in relation to tau species present. In **A** are plotted the size of PSD95 puncta in AD and control cases depending on the presence of tau species. In **B** are plotted the mean signal intensity of postsynaptic terminals in AD and control cases in relation to the presence of tau species. Boxplots show quartiles and medians calculated from each image stack. Data points refer to case means (females, triangles; males, circles). Analysis was with linear mixed effects models including diagnostic group, sex and tau specie presence. *p<0.05 in post-hoc emmeans test after linear mixed effects model. Abbreviations: a.u., arbitrary units. Scale bars represent 20 μm.

**Supplementary Figure 3.**
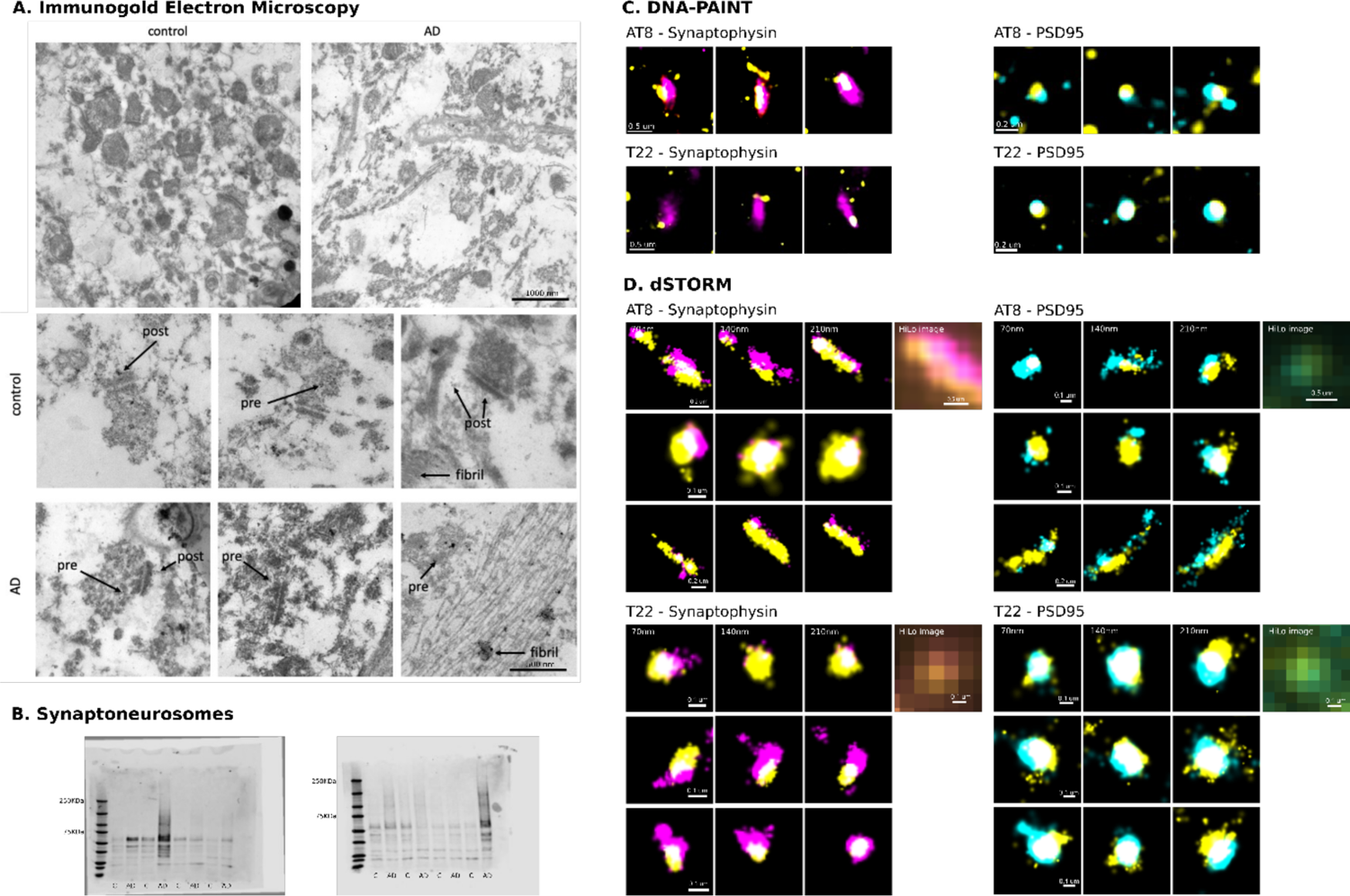
Confirmation of AD synaptic tau aggregates. **A** Immuno-electron microscopy with gold-conjugated secondary antibodies (black dots) shows T22 at pre- and postsynaptic terminals. Scale bar top = 1000nm bottom = 500nm. **B** Full western blots from synaptoneurosomes isolated from 8 control and 8 AD cases. **C** Nine more examples of DNA-PAINT array tomography 70nm-thick sections confirm tau (yellow) and synaptophysin (magenta) or PSD95 (cyan) colocalization. **D** Nine more examples of dSTORM array tomography 70nm-thick consecutive sections confirm tau (yellow) and synaptophysin (magenta) or PSD95 (cyan) colocalization after reconstruction of HiLo images.

**Supplementary Figure 4.**
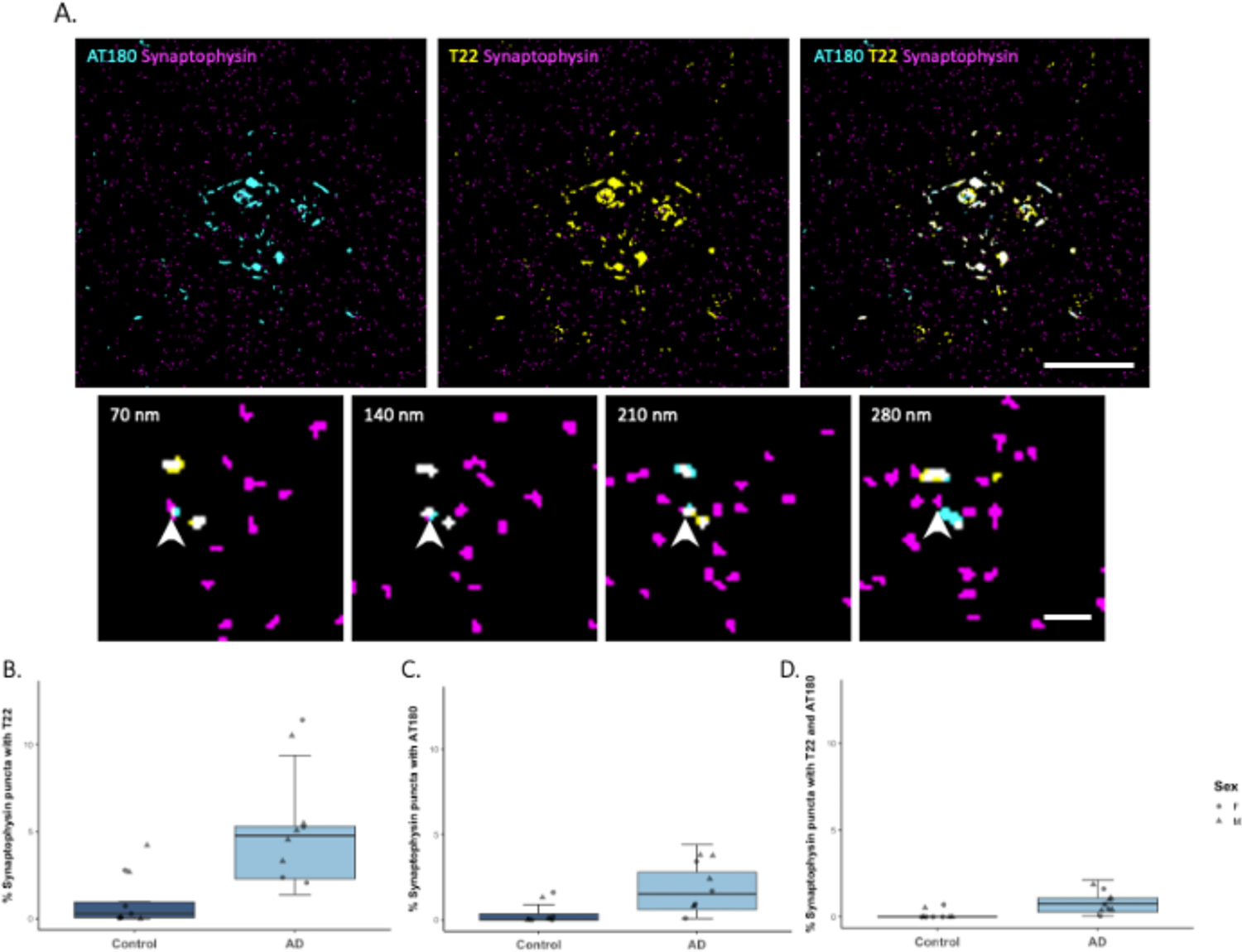
Oligomeric tau is more prevalent than an early phosphoepitope in presynaptic terminals in AD cases. **A** At top, maximum intensity projection of a representative segmented image from array tomography showing presynaptic terminals (magenta), ptau231 (AT180, cyan) and/or oligomeric tau (T22, yellow). The inset below shows four consecutive 70nm thick sections where a presynaptic terminal containing both tau species is pointed by an arrowhead. **B** Percent of presynaptic terminals containing oligomeric tau (T22). **C** Percent of presynaptic terminals containing ptau231 (AT180 clone). **D** Percent of presynaptic terminals containing both oligomeric and ptau231. Boxplots show quartiles and medians calculated from each image stack. Data points refer to case means (females, circles; males, triangles). Analysis was with linear mixed effects models including diagnostic group and sex. Scale bars represent 20 μm and 2 μm.

**Supplementary Figure 5.**
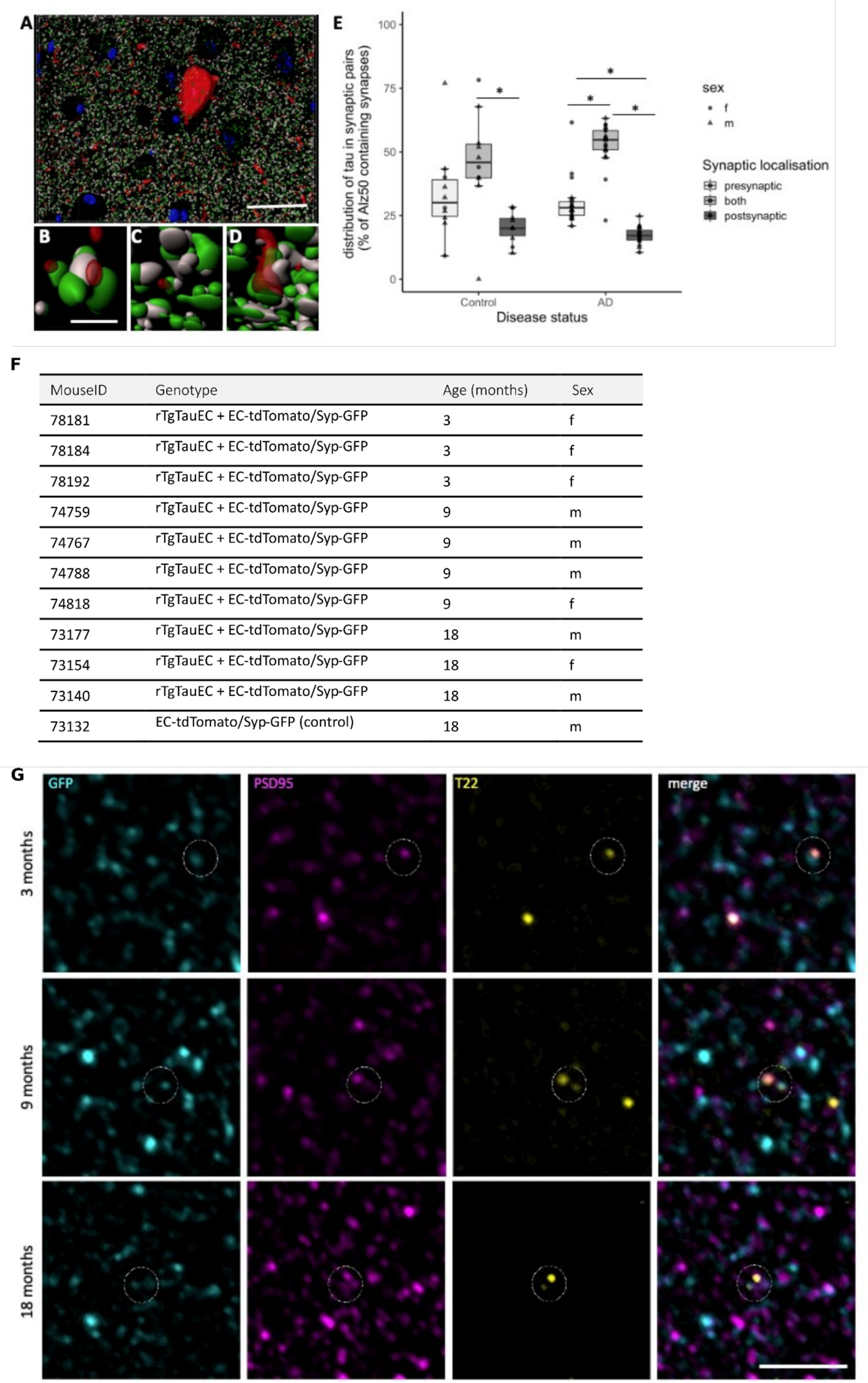
Tau distribution at synaptic terminals. Panel **A** Shows a 3D reconstruction made in Imaris of array tomography images of misfolded tau (Alz50, red), presynaptic terminals (grey), post synaptic terminals (green) and DAPI (blue). We observe tau accumulation in presynaptic terminals (**B**), postsynaptic terminals (**C**) and both sides of synaptic pairs (**D**). Quantification of synaptic pairs containing tau (**E**) reveals most tau accumulation differs between presynaptic only, both sides of the pair, and postsynaptic only (ANOVA after linear model F=62.93, p<0.0001), asterisks represent post-hoc Tukey corrected tests p<0.05. Array tomography imaging of mice expressing P301L mutant human tau and GFP tagged synaptophysin (**F**) in entorhinal cortex stained for GFP, PSD95, and T22 (**G**) show that oligomeric tau can spread from pre to post synapses at 3 ages studied. Scale bars represent 20 μm In A, 1 μm in B-D, 5 μm in F.

**Supplementary Table 1.**
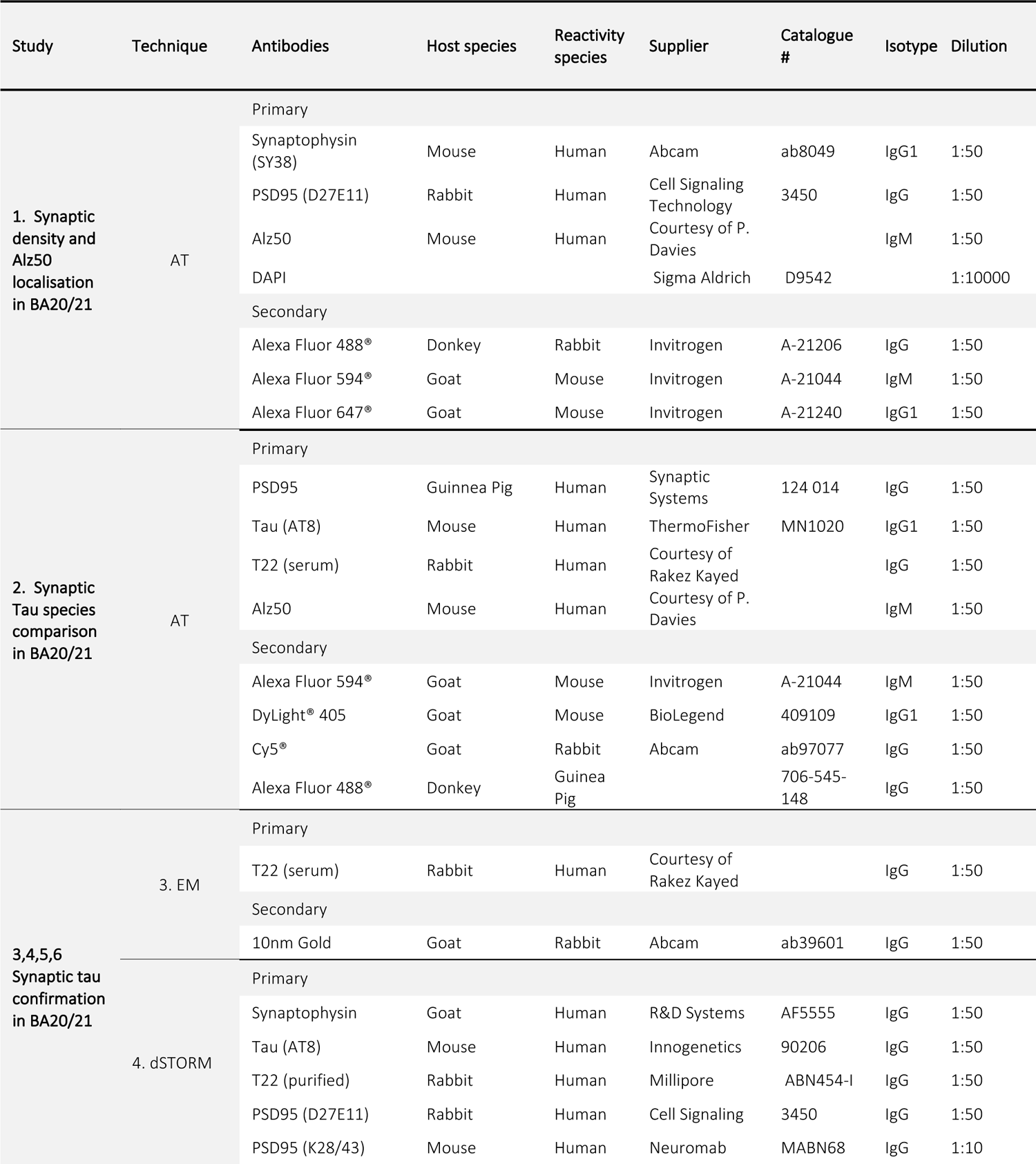
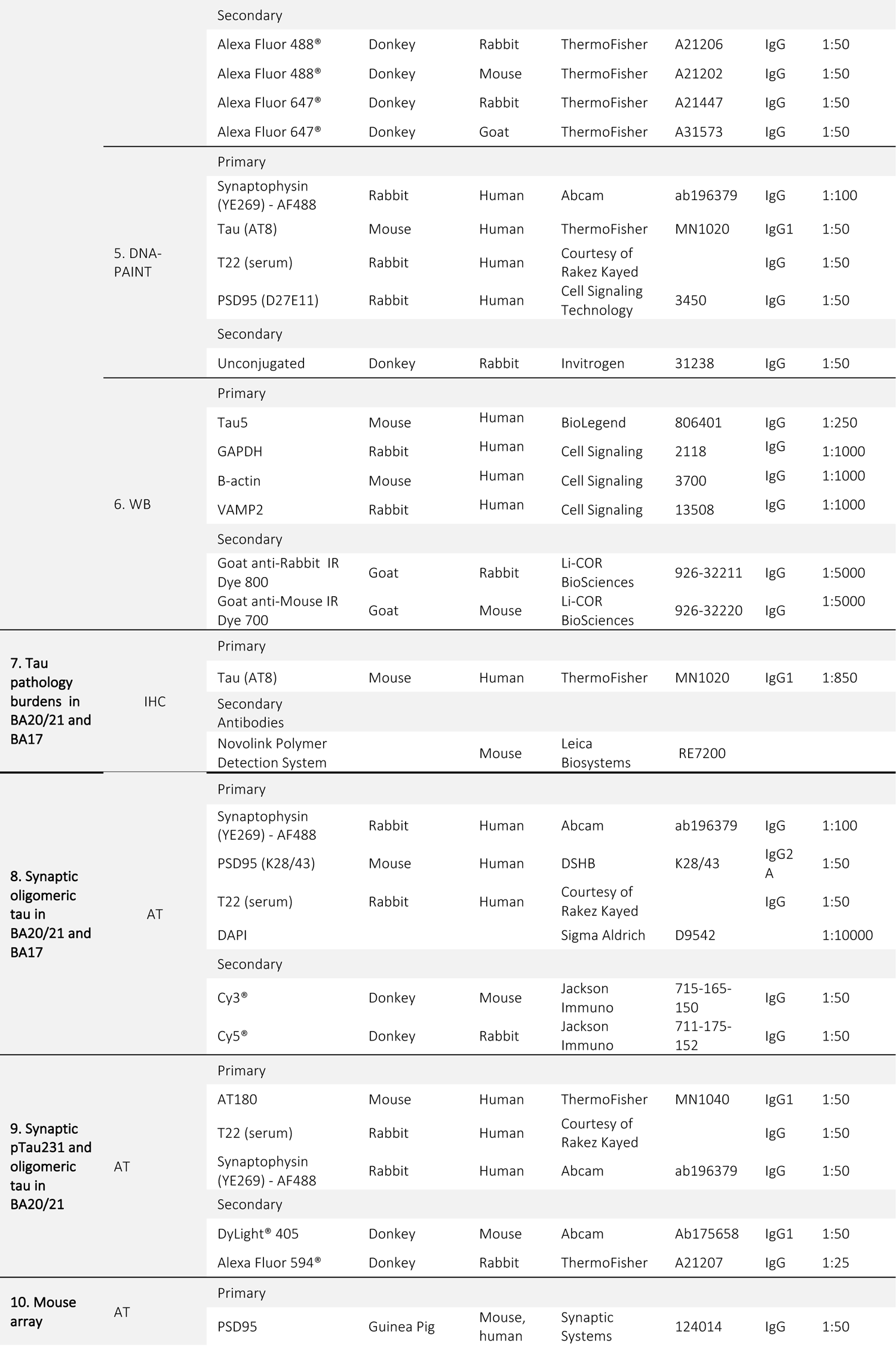
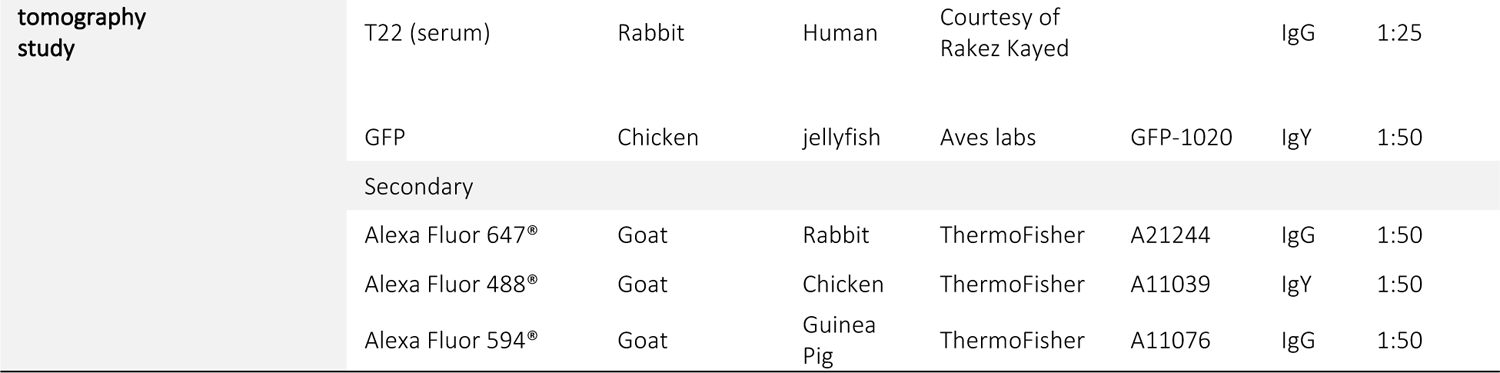
Antibody combinations by experiment. AT: Array tomography microscopy. EM: Immunogold electron microscopy. dSTORM: Direct Stochastic Optical Reconstruction Microscopy. WB: Western blot. IHC: Immunohistochemistry.

